# Long-Term Mortality with Contemporary Multivessel PCI Versus CABG in Patients with Diabetes and Multivessel Disease: A Population-Based Retrospective Cohort Study

**DOI:** 10.64898/2026.09.04.26362312

**Authors:** Xiang Xiao, Anamaria Savu, Kevin R. Bainey, Dean Eurich, Roopinder K. Sandhu, Padma Kaul

## Abstract

**Background:** Coronary artery bypass grafting (CABG) is often preferred over percutaneous coronary intervention (PCI) in patients with diabetes mellitus (DM) and multivessel coronary artery disease (MVD), but whether it confers a long-term survival advantage remains unclear. Moreover, whether anatomic complexity modulates this relationship is unknown.

**Methods:** We included 5,615 patients with diabetes and MVD who underwent revascularization with contemporary drug-eluting stents (PCI-DES; n=3,410) or CABG (n=2,205) in Alberta, Canada, between 2009 and 2019. The primary outcome was 5-year all-cause mortality. Secondary outcomes were 5-year major adverse cardiovascular events (MACE) and 10-year mortality and MACE in an earlier-era subcohort of 2,103 patients. Inverse probability of treatment weighting with Cox proportional hazards models was used. Interactions between revascularization, Duke Jeopardy Score (DJS), and clinical presentation (stable angina, unstable angina/NSTEMI, or STEMI) were examined.

**Results:** Five-year all-cause mortality did not differ between PCI-DES and CABG (HR, 1.14; [95% CI,0.99-1.31]; P=0.08). However, PCI-DES was associated with a higher 5-year MACE (HR, 1.23; [95% CI,1.10-1.38]; P=0.0004), but lower 10-year all-cause mortality compared to CABG (HR,0.85; [95% CI,0.72-0.99]; P=0.04) with no difference in MACE (HR, 0.97; 95 % CI: 0.85-1.11; P=0.68). Anatomic complexity modified these findings, with lower 5-year mortality after CABG only at higher DJS (P_interaction_=0.003), and lower 10-year mortality after PCI-DES only at lower DJS (P_interaction_<0.0001).

**Conclusions:** In patients with diabetes and MVD, 5-year all-cause mortality did not differ significantly between groups, although PCI-DES was associated with higher 5-year MACE rates. At 10 years, multivessel PCI-DES was associated with lower 10-year mortality and no difference in MACE. Anatomic complexity modified the effects, favouring CABG at higher DJS and PCI-DES at lower DJS. Our findings should be considered when planning revascularization in diabetes with MVD.

## Introduction

For patients with both diabetes mellitus (DM) and multivessel coronary artery disease (MVD), North American and European guidelines for both acute and chronic coronary syndromes have historically recommended coronary artery bypass grafting (CABG) over percutaneous coronary intervention (PCI) as the preferred revascularization strategy.^1–5^ This recommendation is grounded in evidence from landmark randomized controlled trials, such as FREEDOM (Future Revascularization Evaluation in Patients with Diabetes Mellitus: Optimal Management of Multivessel Disease), which demonstrated the benefit of CABG over first-generation drug-eluting stents (DES) in reducing long-term mortality and major adverse cardiovascular events with a median follow-up of 3.8 years.^6^ Extended follow-up of the FREEDOM cohort to a median of 7.5 years subsequently reported lower all-cause mortality after CABG.^7^ However, findings from more recent studies examining survival outcomes of PCI-DES and CABG in patients with diabetes and MVD have been less consistent. A previous cohort study showed PCI-DES was associated with a higher risk of mortality,^8^ whereas others reported no statistically significant difference in survival between the two strategies.^9^ These divergent results suggest that the comparative effectiveness of multivessel PCI and CABG may depend on patient characteristics, disease anatomy, and contemporary practice patterns. Moreover, evidence in patients with diabetes suggests that mortality differences between PCI-DES and CABG may be attenuated in the newer-generation DES era,^10^ and findings from a cohort specifically limited to patients with diabetes and MVD similarly report comparable long-term mortality with everolimus-eluting stents (EES)-PCI versus CABG, despite differences in non-fatal events.^11^ Of note, data on contemporary long-term survival comparisons beyond 5 years remain sparse.

Accordingly, using a large contemporary population-based cohort of patients with diabetes and MVD who underwent revascularization, we compared long-term clinical outcomes between patients treated with PCI using contemporary drug-eluting stents (PCI-DES) and those treated with CABG. Furthermore, we evaluated whether anatomic complexity, as defined by the Duke Jeopardy Score (DJS), and clinical presentation modify the association between revascularization strategy and outcomes.^12^

## Methods

### Patient population

The retrospective cohort population included patients diagnosed with diabetes and MVD who underwent either PCI-DES or CABG within 90 days of index cardiac catheterization (to allow for outpatient CABG and staged multivessel PCI) between January 1, 2009, and March 31, 2019, in Alberta, Canada. This time interval was chosen to identify patients who received at least a second-generation DES (contemporary DES) with adequate long-term follow-up. Based on an established Canadian Chronic Disease Surveillance System definition, patients were considered to have diabetes if it was listed in any diagnosis field of one hospital separation or two physician office visit records within two years.^13^ MVD was defined as stenosis greater than 75% in at least two coronary vessels or the left main coronary artery, with inclusion further restricted to a DJS of 4 or higher where available.^12^

### Inclusion and exclusion criteria

Patients with diabetes and MVD who underwent index cardiac catheterization for chronic coronary syndrome - stable angina (SA), or acute coronary syndrome - unstable angina (UA), non-ST-elevation myocardial infarction (NSTEMI), or ST-elevation myocardial infarction (STEMI) were included. The index catheterization was each patient’s first in the APPROACH registry, with diabetes ascertained on or before that date. We excluded patients who were < 18 years old, did not meet the MVD criteria defined above, had prior revascularization or cardiac or great-vessel intervention (Table S1), were non-Alberta residents, or did not undergo PCI-DES or CABG within 90 days of catheterization. The PCI cohort was restricted to second-generation DES (e.g., Promus, Xience, Resolute, Orsiro). Patients with missing or non-qualifying stent data were excluded.

### Data source and linkage

The APPROACH (Alberta Provincial Project for Outcomes Assessment in Coronary Heart Diseases) registry includes detailed demographic, clinical, and cardiac anatomical data from patients who underwent cardiac catheterization in Alberta. Details of the dataset have been described previously.^14^ In this study, the APPROACH registry was linked to the Alberta Diabetes Registry, the Discharge Abstract Database (DAD), the National Ambulatory Care Reporting System (NACRS), Practitioner Claims, Vital Statistics, the Alberta Health Care Insurance Plan Registry, and the Pharmaceutical Information Network (PIN) using a scrambled unique patient identifier. These administrative databases are commonly used in research and have been validated previously.^15,16^

### Exposure and Baseline Characteristics

The exposure of interest was revascularization within 90 days of index cardiac catheterization, categorized as either PCI-DES or CABG to account for stable patients requiring outpatient revascularization, as is the practice in Alberta. Staged procedures were considered part of the index intervention. Patients receiving PCI-DES and CABG within 90 days of the index procedure were assigned to the CABG group. The date of the final revascularization within the 90-day window was defined as the index exposure date. Baseline characteristics included age, sex, residence status,^17^ ethnicity, smoking status, body mass index, dialysis status, hyperlipidemia, selected comorbidities, extent of coronary artery disease (CAD), clinical presentation of CAD, Charlson Comorbidity Index,^15^ DJS, and the Pampalon Material Deprivation Index.^18,19^ Ethnicity was identified based on the patient’s surname using previously validated algorithms and categorized as South Asian or Chinese.^20–22^ Patients not in either group were categorized as unclassified, predominantly of European ancestry in Alberta.^23^ Covariate selection was based on their clinical relevance and review of existing literature.^24,25^ Comorbidities were identified from DAD, NACRS, and physician claims using ICD-9/10 codes (Table S2). Medication dispensations within six months before and after revascularization were obtained from the PIN database. Missing BMI and DJS were addressed using stochastic regression imputation.^26^ Missing material deprivation index was retained as a separate category. Ethics approval for the study was obtained from the Health Research Ethics Board of the University of Alberta (Pro00137996). As data came from health databases, informed consent was waived.

### Clinical Outcomes

The primary outcome was all-cause mortality at 5 years. Key secondary outcomes were major adverse cardiovascular events (MACE) at 5 years and mortality and MACE at 10 years. Other endpoints were Major Adverse Cardiac and Cerebrovascular Events (MACCE) and the individual components of the composite outcomes. Mortality and MACE at 30 days were examined for comparison purposes. MACE was defined as a composite of all-cause mortality, myocardial infarction (MI), or stroke. MACCE was defined as a composite of all-cause mortality, stroke, MI, or repeat revascularization. Repeat revascularization was defined as any revascularization procedure performed after the final revascularization within the 90-day window, and ≥6 weeks after PCI-DES or ≥4 weeks after CABG. Only the first qualifying event was considered. Deaths were identified from Vital Statistics databases. MI and stroke were identified using the primary diagnosis field in the hospitalization and emergency department visit records, excluding events that occurred during the index hospitalization. Cardiovascular deaths were identified from the Vital Statistics using ICD-10 codes (I00-I99). All-cause mortality at 10 years was examined among patients with a catheterization date before December 31^st^, 2013, to allow at least 10 years of potential follow-up. As a supportive analysis to assess whether long-term all-cause mortality patterns were consistent for cardiovascular deaths, 10-year cardiovascular mortality was also examined in this restricted cohort.

Patients were followed from index revascularization until the earliest occurrence of the outcome of interest, death, migration out of province, or the end of the study period (December 31^st^, 2023), whichever came first. Individuals who did not experience the outcome of interest were censored at the end of follow-up. The corresponding ICD-10 codes for all outcomes are provided in Table S3.

### Statistical analysis

Baseline characteristics were summarized by revascularization using counts and percentages for categorical variables and means with standard deviations or medians with interquartile ranges for continuous variables. Group comparisons were conducted using the Chi-square test for categorical variables, and the t-test or Wilcoxon rank-sum test for continuous variables. To control for confounding, we created a pseudo-population using inverse probability of treatment weighting (IPTW). Propensity scores were estimated using a multivariable logistic regression model including the following baseline covariates: age, sex, residence status, smoking, dialysis, hyperlipidemia, ethnicity, Pampalon Material Deprivation Index, hypertension, Charlson Comorbidity Index, clinical presentation of CAD, and BMI. Balance after weighting was assessed using standardized mean differences, with a value <0.1 indicating adequate balance.^27^ Event rates were reported as crude proportions and as incidence rates per 100 person-years. Survival was estimated using the Kaplan–Meier (K-M) method and compared between groups with the log-rank test. Cumulative incidence functions (CIF) accounted for the competing risk of death for non-fatal outcomes. IPTW-weighted Cox proportional hazards models with robust variance estimators were used to estimate adjusted hazard ratios (HRs) with 95% confidence intervals for all-cause mortality, MACE, and MACCE.^28^ For individual non-fatal outcomes, cause-specific hazard models were fitted using the same IPTW weights. The proportional hazards assumption was tested using time-by-treatment interaction terms and was not violated for the outcomes.

To evaluate whether treatment effects varied by anatomic complexity, an interaction term between revascularization and DJS was included in the IPTW-weighted Cox models. An interaction with clinical presentation was also examined and is reported in the Supplement. We conducted a sensitivity analysis restricting the cohort to patients with a catheterization date before December 31st, 2013, to compare 5-year and 10-year outcomes within the same population. To assess whether effect modification by DJS was driven by left main disease, the model including interaction between revascularization and DJS was refitted, excluding patients with left main at both 5 and 10 years. All statistical analyses were performed using SAS software, version 9.4 (SAS Institute, Cary, NC). A two-sided p-value < 0.05 was considered statistically significant. Reporting of this study adhered to the Strengthening the Reporting of Observational Studies in Epidemiology (STROBE) statement.

## Results

Among 5,615 patients with diabetes and MVD who underwent revascularization within 90 days of index catheterization between January 1, 2009, and March 31, 2019, 3,410 (60.7%) received PCI-DES and 2,205 (39.3%) received CABG (Figure 1). The proportion of CABG rather than PCI-DES increased with DJS (11.1% with DJS of 4 to 78.0 % with DJS of 12). The overall median follow-up for the entire cohort was 8.6 years (interquartile range: 6.7, 11.5) (PCI-DES: 9.0 (6.9, 12.0) vs. CABG: 8.4 (6.6, 11.0); P<0.0001). A total of 11 patients (7 in PCI-DES; 4 in CABG) migrated out of the province during the follow-up. All migrations occurred within the first 5 years.

**Figure 1.**
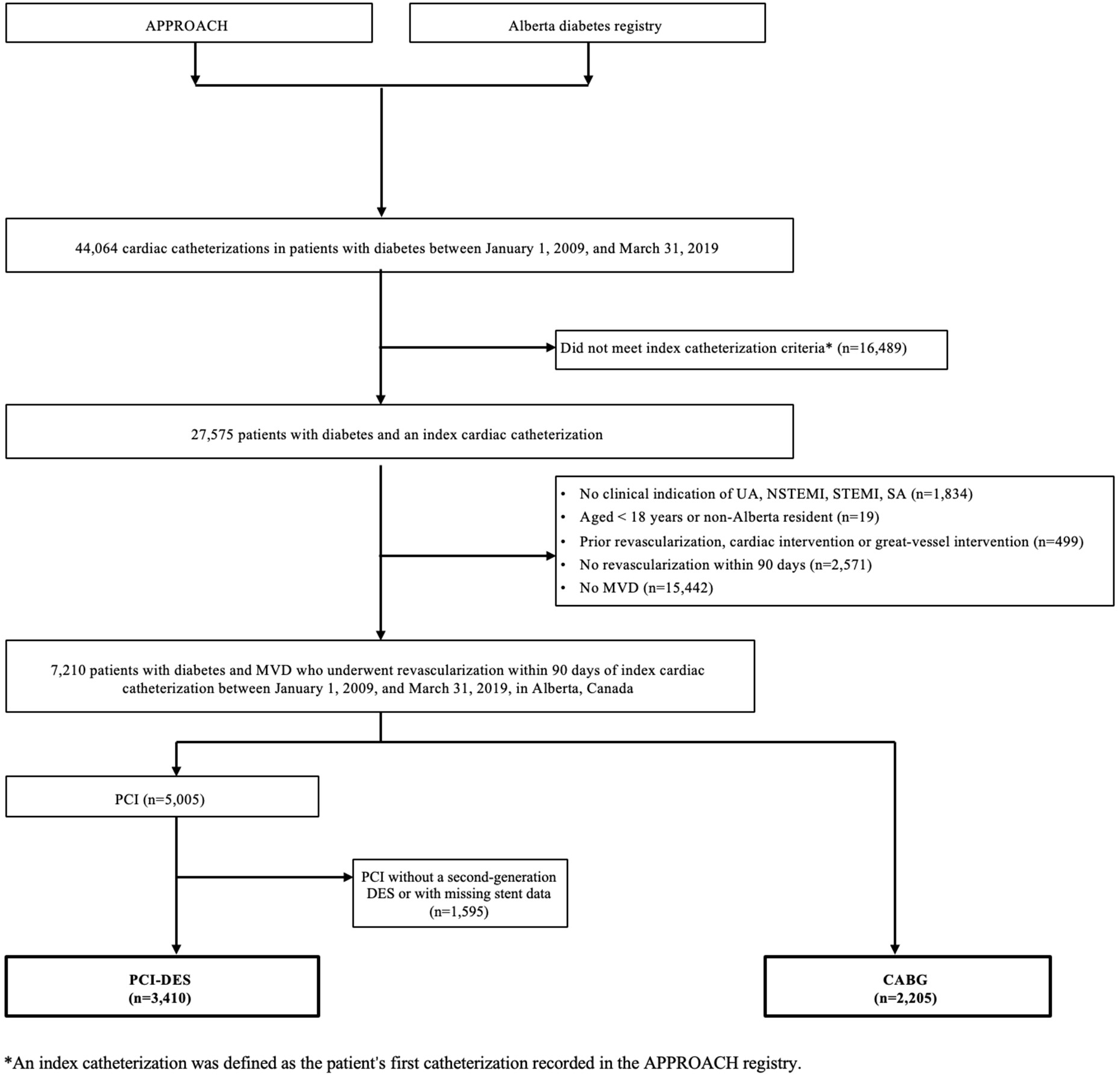
Flow diagram of the study cohort.

### Baseline characteristics by revascularization

Compared to patients who received CABG, those who received PCI-DES were more likely to be female, younger, present with STEMI, obese, and less likely to be smokers. However, the CABG group had a substantially higher comorbidity burden. The prevalence of most comorbidities, including hypertension, heart failure, cerebrovascular disease, and peripheral vascular disease, was significantly higher among patients who underwent CABG (Table 1). Additionally, the CABG group had a significantly greater burden of coronary disease with a much higher prevalence of left main disease and a higher median DJS. Medication use by revascularization is provided in Table S4. BMI and DJS had 14.0 % and 5.2 % missing, respectively, as shown in Table 1.

**Table 1.**
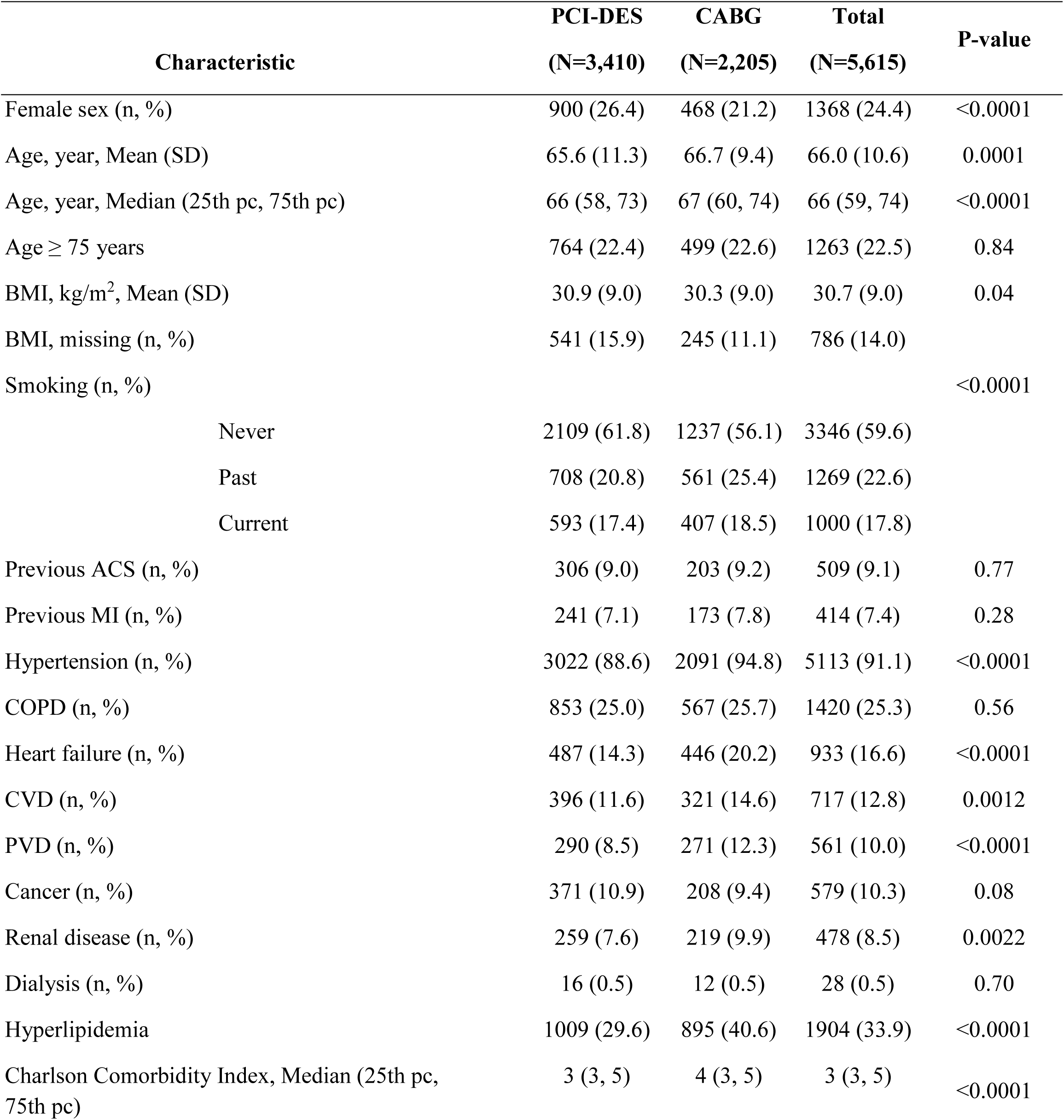
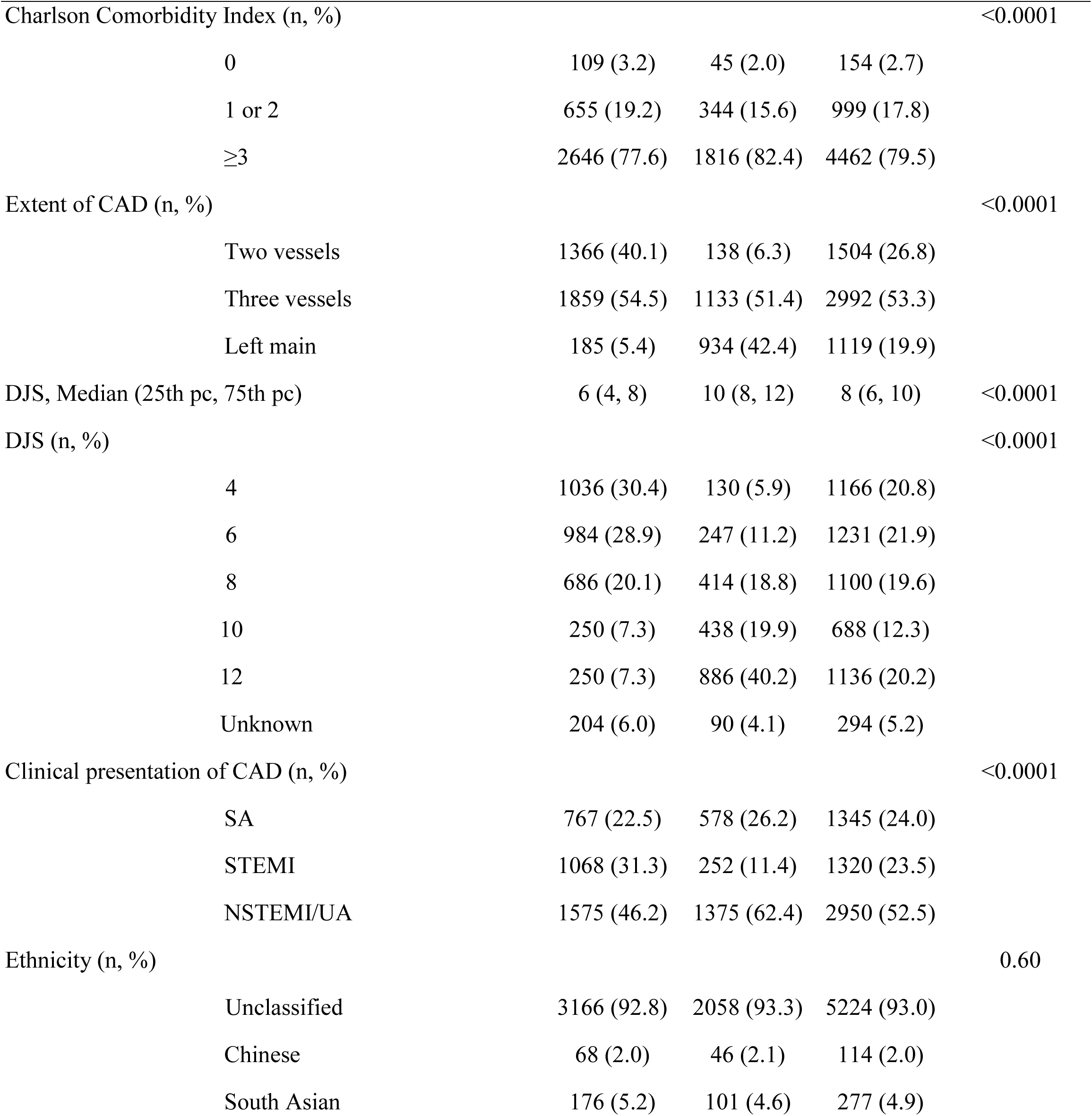
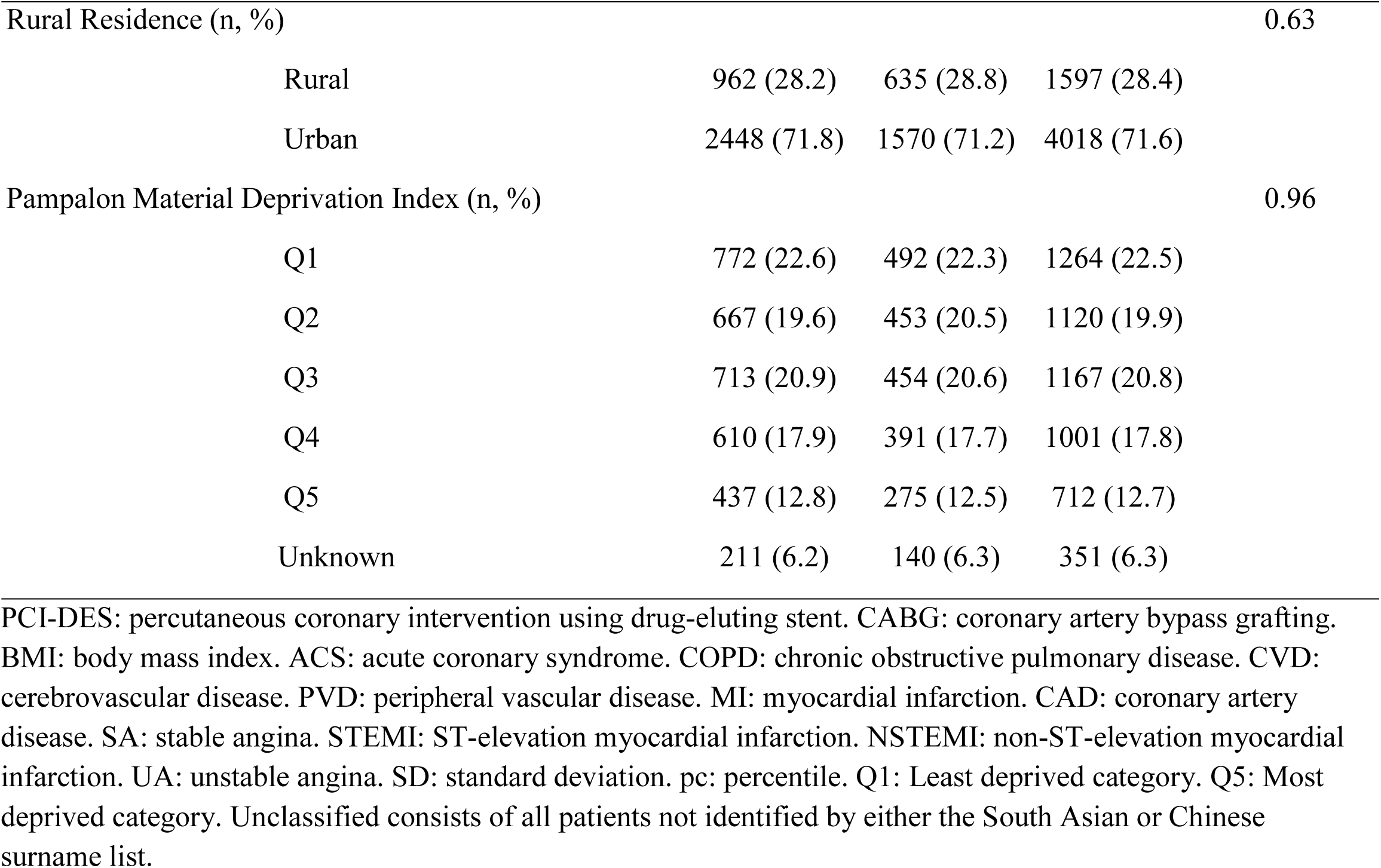
Patient baseline characteristics at catheterization.

| Characteristic |  | PCI-DES<br>(N=3,410) | CABG<br>(N=2,205) | Total<br>(N=5,615) | P-value |
| --- | --- | --- | --- | --- | --- |
| Female sex (n, %) |  | 900 (26.4) | 468 (21.2) | 1368 (24.4) | <0.0001 |
| Age, year, Mean (SD) |  | 65.6 (11.3) | 66.7 (9.4) | 66.0 (10.6) | 0.0001 |
| Age, year, Median (25th pc, 75th pc) |  | 66 (58, 73) | 67 (60, 74) | 66 (59, 74) | <0.0001 |
| Age ≥ 75 years | | 764 (22.4) | 499 (22.6) | 1263 (22.5) | 0.84 |
| BMI, kg/m<sup>2</sup>, Mean (SD) |  | 30.9 (9.0) | 30.3 (9.0) | 30.7 (9.0) | 0.04 |
| BMI, missing (n, %) |  | 541 (15.9) | 245 (11.1) | 786 (14.0) |  |
| Smoking (n, %) |  |  |  |  | <0.0001 |
|  | Never | 2109 (61.8) | 1237 (56.1) | 3346 (59.6) |  |
|  | Past | 708 (20.8) | 561 (25.4) | 1269 (22.6) |  |
|  | Current | 593 (17.4) | 407 (18.5) | 1000 (17.8) |  |
| Previous ACS (n, %) |  | 306 (9.0) | 203 (9.2) | 509 (9.1) | 0.77 |
| Previous MI (n, %) |  | 241 (7.1) | 173 (7.8) | 414 (7.4) | 0.28 |
| Hypertension (n, %) |  | 3022 (88.6) | 2091 (94.8) | 5113 (91.1) | <0.0001 |
| COPD (n, %) |  | 853 (25.0) | 567 (25.7) | 1420 (25.3) | 0.56 |
| Heart failure (n, %) |  | 487 (14.3) | 446 (20.2) | 933 (16.6) | <0.0001 |
| CVD (n, %) |  | 396 (11.6) | 321 (14.6) | 717 (12.8) | 0.0012 |
| PVD (n, %) |  | 290 (8.5) | 271 (12.3) | 561 (10.0) | <0.0001 |
| Cancer (n, %) |  | 371 (10.9) | 208 (9.4) | 579 (10.3) | 0.08 |
| Renal disease (n, %) |  | 259 (7.6) | 219 (9.9) | 478 (8.5) | 0.0022 |
| Dialysis (n, %) |  | 16 (0.5) | 12 (0.5) | 28 (0.5) | 0.70 |
| Hyperlipidemia |  | 1009 (29.6) | 895 (40.6) | 1904 (33.9) | <0.0001 |
| Charlson Comorbidity Index, Median (25th pc, 75th pc) |  | 3 (3, 5) | 4 (3, 5) | 3 (3, 5) | <0.0001 |

| Characteristic | PCI-DES<br>(N=3,410) | CABG<br>(N=2,205) | Total<br>(N=5,615) | P-value |
| --- | --- | --- | --- | --- |
| Charlson Comorbidity Index (n, %) |  |  |  | <0.0001 |
| 0 | 109 (3.2) | 45 (2.0) | 154 (2.7) |  |
| 1 or 2 | 655 (19.2) | 344 (15.6) | 999 (17.8) |  |
| ≥3 | 2646 (77.6) | 1816 (82.4) | 4462 (79.5) |  |
| Extent of CAD (n, %) |  |  |  | <0.0001 |
| Two vessels | 1366 (40.1) | 138 (6.3) | 1504 (26.8) |  |
| Three vessels | 1859 (54.5) | 1133 (51.4) | 2992 (53.3) |  |
| Left main | 185 (5.4) | 934 (42.4) | 1119 (19.9) |  |
| DJS, Median (25th pc, 75th pc) | 6 (4, 8) | 10 (8, 12) | 8 (6, 10) | <0.0001 |
| DJS (n, %) |  |  |  | <0.0001 |
| 4 | 1036 (30.4) | 130 (5.9) | 1166 (20.8) |  |
| 6 | 984 (28.9) | 247 (11.2) | 1231 (21.9) |  |
| 8 | 686 (20.1) | 414 (18.8) | 1100 (19.6) |  |
| 10 | 250 (7.3) | 438 (19.9) | 688 (12.3) |  |
| 12 | 250 (7.3) | 886 (40.2) | 1136 (20.2) |  |
| Unknown | 204 (6.0) | 90 (4.1) | 294 (5.2) |  |
| Clinical presentation of CAD (n, %) |  |  |  | <0.0001 |
| SA | 767 (22.5) | 578 (26.2) | 1345 (24.0) |  |
| STEMI | 1068 (31.3) | 252 (11.4) | 1320 (23.5) |  |
| NSTEMI/UA | 1575 (46.2) | 1375 (62.4) | 2950 (52.5) |  |
| Ethnicity (n, %) |  |  |  | 0.60 |
| Unclassified | 3166 (92.8) | 2058 (93.3) | 5224 (93.0) |  |
| Chinese | 68 (2.0) | 46 (2.1) | 114 (2.0) |  |
| South Asian | 176 (5.2) | 101 (4.6) | 277 (4.9) |  |
| Rural Residence (n, %) |  |  |  | 0.63 |
| Rural | 962 (28.2) | 635 (28.8) | 1597 (28.4) |  |
| Urban | 2448 (71.8) | 1570 (71.2) | 4018 (71.6) |  |
| Pampalon Material Deprivation Index (n, %) |  |  |  | 0.96 |
| Q1 | 772 (22.6) | 492 (22.3) | 1264 (22.5) |  |
| Q2 | 667 (19.6) | 453 (20.5) | 1120 (19.9) |  |
| Q3 | 713 (20.9) | 454 (20.6) | 1167 (20.8) |  |
| Q4 | 610 (17.9) | 391 (17.7) | 1001 (17.8) |  |
| Q5 | 437 (12.8) | 275 (12.5) | 712 (12.7) |  |
| Unknown | 211 (6.2) | 140 (6.3) | 351 (6.3) |  |

PCI-DES: percutaneous coronary intervention using drug-eluting stent. CABG: coronary artery bypass grafting. BMI: body mass index. ACS: acute coronary syndrome. COPD: chronic obstructive pulmonary disease. CVD: cerebrovascular disease. PVD: peripheral vascular disease. MI: myocardial infarction. CAD: coronary artery disease. SA: stable angina. STEMI: ST-elevation myocardial infarction. NSTEMI: non-ST-elevation myocardial infarction. UA: unstable angina. SD: standard deviation. pc: percentile. Q1: Least deprived category. Q5: Most deprived category. Unclassified consists of all patients not identified by either the South Asian or Chinese surname list.

### Outcomes at 30 days and 5 years

All-cause mortality at 30 days following the index revascularization was 2.9 % after PCI-DES and 1.8 % after CABG (P_log-rank_=0.01). This early difference was driven predominantly by patients with STEMI (6.0% vs 1.2%, P_log-rank_=0.002), whereas mortality in non-STEMI patients did not differ (1.5% vs 1.9%, P_log-rank_=0.23). As seen in Figure 2, the 5-year mortality was 16.3% vs 15.6% (P_log-rank_ = 0.38). PCI-DES had a higher risk of MACE at 30 days (4.5% vs. 3.0% P_log-rank_=0.01). This was also seen at 5 years (27.4% vs. 24.3%, P_log-rank_=0.01) (Figure 2). MACCE was also higher in the PCI-DES group compared to the CABG group (40.7% vs 30.1%, P_log-rank_<0.0001). Among the components of MACCE, the difference between PCI-DES and CABG was most pronounced for repeat revascularization, which was substantially higher in the PCI-DES group (Gray’s test P<0.0001). The risk of MI was also significantly higher for PCI-DES at 5 years, but no difference was observed in cardiovascular death. In contrast, increased risk of stroke was observed in CABG at 5 years (Gray’s test, P=0.04) (Figure S1). The direction of these differences was unchanged when expressed as incidence rates per 100 person-years, accounting for the shorter follow-up in the CABG group.

**Figure 2.**
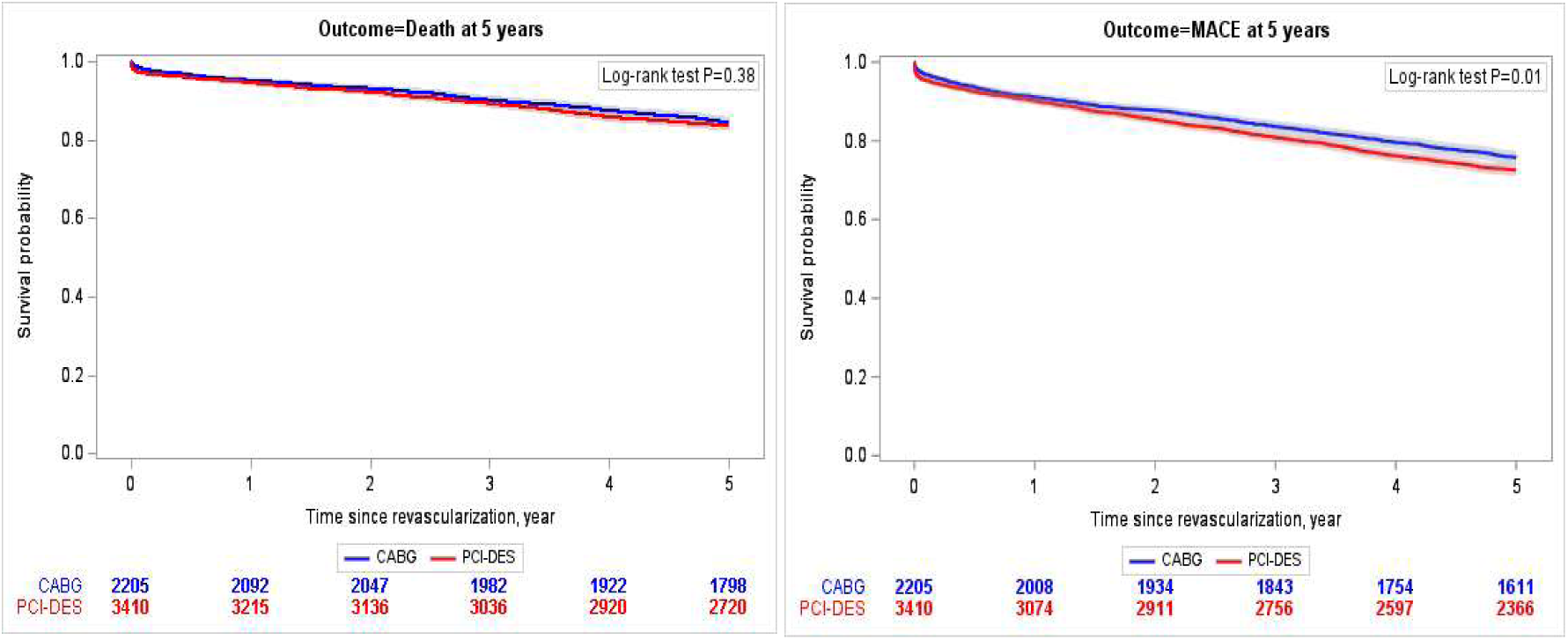
Kaplan-Meier curves for all-cause mortality and MACE at 5 years, comparing PCI-DES versus CABG.

After IPTW adjustment, the standardized mean differences of the baseline characteristics of interest between PCI-DES and CABG were all < 0.10 (Figure S2). At five years, the risk of death was comparable between PCI-DES and CABG (HR, 1.14; [95% CI, 0.99-1.31]; P=0.08). The PCI-DES group showed a significantly higher risk of MACE compared to the CABG group (HR,1.23; [95% CI,1.10-1.38]; P=0.0004) (Figure 3). The PCI-DES group had a higher risk of MACCE than the CABG group (HR, 1.58; [95% CI, 1.43-1.74]; P<0.0001). PCI-DES was also associated with a higher risk of MI and repeat revascularization compared to CABG. However, no difference was observed in cardiovascular (CV) death (Figure S3).

**Figure 3.**
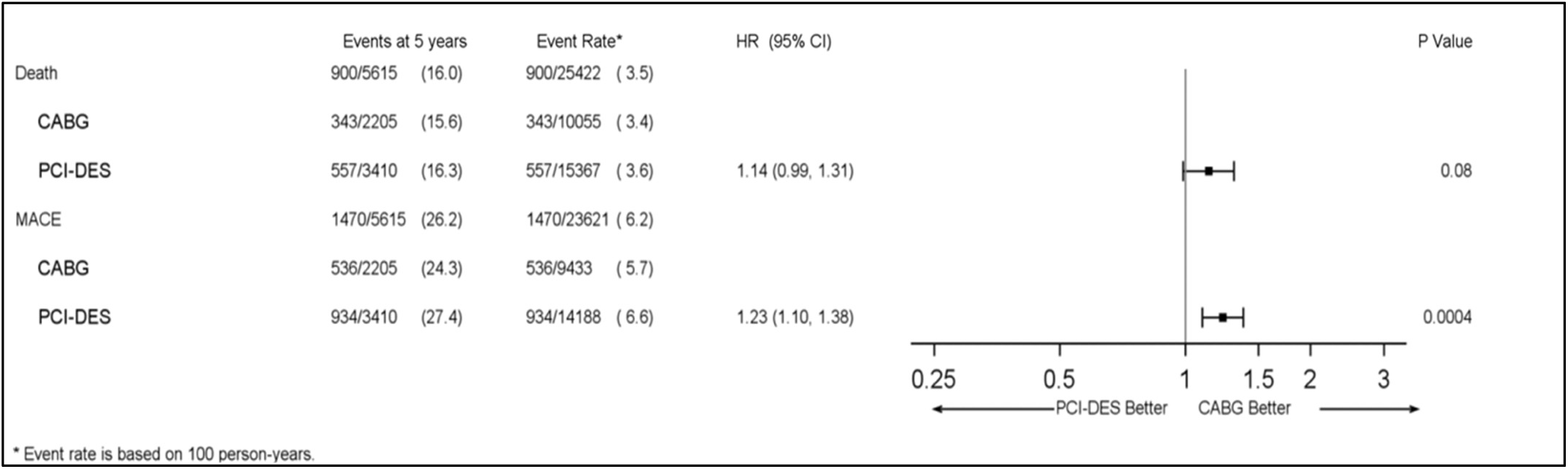
Forest plot of hazard ratios for 5-year all-cause mortality and MACE from IPTW-weighted Cox models.

### Outcomes by anatomical complexity and clinical presentation

At 5 years, the association between revascularization and the risk of death varied according to DJS (P_interaction_=0.003; Figure 4). We observed no statistically significant differences in the risk of death for patients with low-to-moderate anatomic complexity (DJS 4-8). In contrast, among patients with high-complexity disease, PCI-DES was associated with a substantially increased risk of death. The adjusted HR was 1.96 (95% CI,1.48, 2.58; P<0.0001] for a DJS of 10 and 2.56 [95% CI,1.99, 3.30; P<0.0001] for a score of 12. For other outcomes, a similar trend was observed for MACE (P_interaction_=0.003) and CV death (P_interaction_ < 0.0001). However, PCI-DES was associated with higher risks of MACCE and MI across all DJS levels (P_interaction_=0.005 and 0.008). A similar pattern was observed for repeat revascularization, although effect modification was not statistically significant (P_interaction_=0.07) (Figure S4). After excluding 1,119 patients with left main disease, 95.8% of whom had a DJS greater than or equal to 10, the interaction was not statistically significant for MACE (P_interaction_=0.28), MACCE (P_interaction_=0.19), or death (P _interaction_=0.78). The interaction between clinical presentation and revascularization was not statistically significant (P_interaction_=0.07), and stratified estimates are provided in Figures S5 and S6.

**Figure 4.**
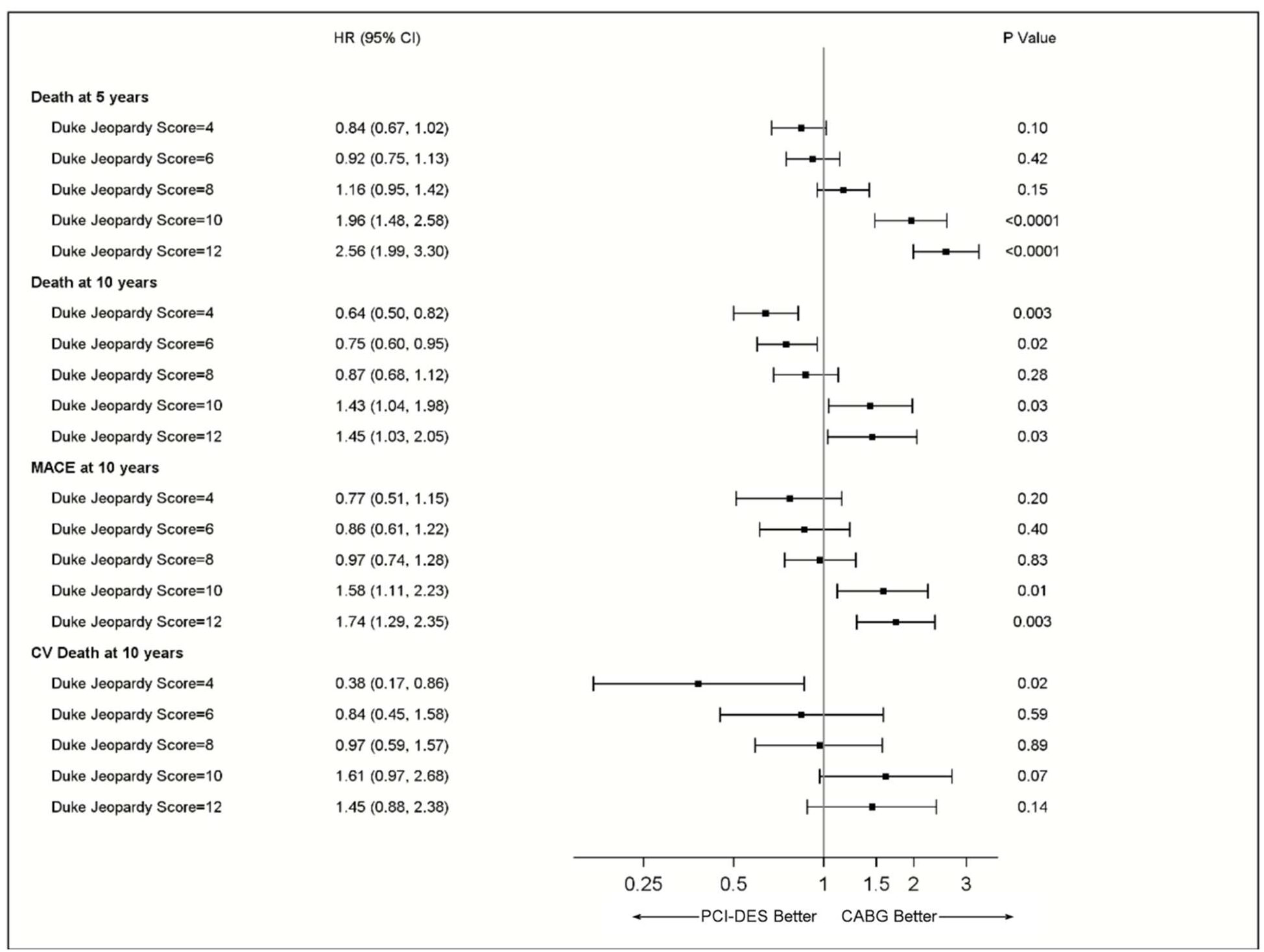
Forest plot of IPTW-weighted Cox model hazard ratios for 5-year and 10-year all-cause mortality, and 10-year MACE and cardiovascular mortality across DJS levels.

### Outcomes at 10 years

A total of 2,103 patients who underwent cardiac catheterization before December 31^st^, 2013, were eligible for potential 10-year follow-up. Of these, 948 (45.1%) had CABG, and 1,155 (54.9%) had PCI-DES. In this subgroup, mortality was higher after CABG compared to PCI-DES (36.2% vs 30.3%; P_log-rank_=0.001), whereas MACE did not differ (46.6 % vs 44.7 %; P_log-rank_=0.43) (Figure 5). The adjusted hazard ratio for all-cause mortality at 10 years compared PCI-DES with CABG was 0.85 [95 % CI: 0.72-0.99]; P=0.04, MACE was 0.97 [95 % CI: 0.85-1.11]; P=0.68, and cardiovascular mortality was 0.78 (95 % CI: 0.62-0.99; P=0.04).

**Figure 5.**
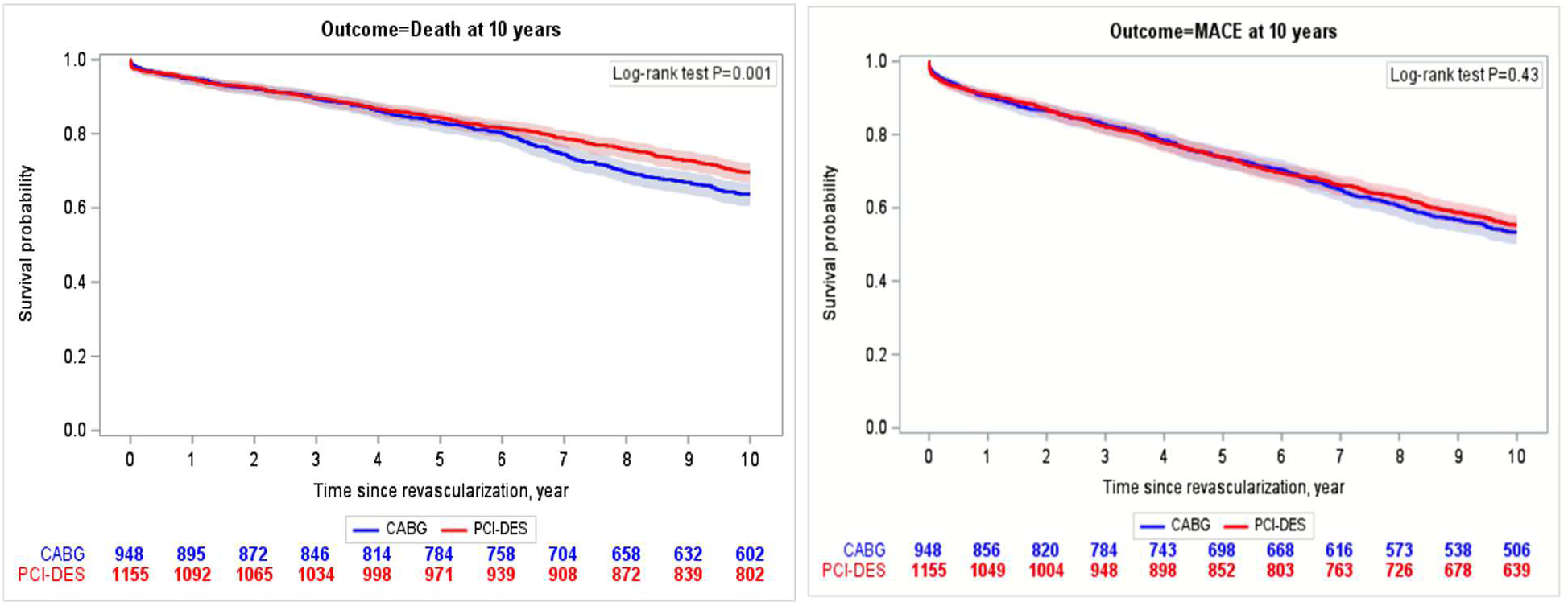
Ten-year Kaplan-Meier curves of all-cause mortality and MACE comparing PCI-DES versus CABG (N=2,103).

At 10 years, DJS modified the effect of revascularization on all-cause mortality (P_interaction_ <0.0001), MACE (P_interaction_=0.001), and cardiovascular mortality (P_interaction_=0.03) (Figure 4). PCI-DES was associated with lower all-cause mortality at a DJS of 4 (HR, 0.64; [95% CI, 0.50-0.82]; P=0.003) and 6 (HR, 0.75; [95% CI, 0.60-0.95]; P=0.02), and with higher mortality at a DJS of 10 (HR,1.43; [95% CI, 1.04-1.98]; P=0.03) and 12 (HR,1.45; [95% CI,1.03-2.05]; P=0.03). MACE differed only at the two highest scores (DJS 10: HR,1.58; [95% CI,1.11-2.23]; P=0.01; DJS 12: HR,1.74; [95% CI,1.29-2.35]; P=0.003), and cardiovascular mortality only at a DJS of 4 (HR, 0.38; [95% CI, 0.17-0.86]; P=0.02). Importantly, when repeated in the subcohort, the 5-year DJS gradient retained the same direction (Table S5). After excluding 455 patients with left main disease, the interaction between revascularization and DJS at 10 years remained statistically significant for all-cause mortality (P_interaction_=0.04), MACE (P_interaction_=0.03), and cardiovascular mortality (P_interaction_=0.04).

## Discussion

In this large, population-based cohort of patients with diabetes and MVD who underwent PCI-DES or CABG, 5-year mortality did not differ statistically between PCI-DES and CABG. In the restricted 10-year cohort, PCI-DES was associated with lower mortality. Moreover, PCI-DES was associated with higher 5-year risks of MACE and MACCE. Relative treatment effects varied by coronary anatomical complexity, with CABG tending to be favored for 5-year survival at higher DJS only, and PCI-DES tending to be favored for 10-year survival at lower DJS.

The absence of a statistically significant difference in 5-year all-cause mortality is consistent with previous studies showing that survival differences between PCI-DES and CABG at 5 years were comparable. In the diabetes subgroup of SYNTAX, 5-year all-cause mortality did not differ significantly between PCI-DES and CABG (P=0.065),^29^ while FREEDOM reported a higher 5-year all-cause mortality after PCI-DES than after CABG (16.3% vs 10.9%; P=0.049); the result was borderline significant, with a median of 3.8 years.^6^ A more recent observational study of 418 patients with diabetes and multivessel disease, fractional flow reserve (FFR)-guided multivessel PCI-DES showed similar 5-year all-cause mortality compared to CABG.^30^ Our findings are concordant with these contemporary reports of PCI-DES. Plausible explanations include lower case-fatality rates after PCI ischemic events with contemporary DES technology (e.g., thinner struts with advanced polymer-eluting platforms^31,32^ and advances in secondary preventive therapies (prolonged DAPT, SGLT-2 inhibitors, GLP-1 agonists, etc.).^33,34^ We acknowledge a slightly higher excess 30-day mortality after PCI-DES, but this was seen only among STEMI patients. Although the interaction between clinical presentation and revascularization was not statistically significant, stratified estimates suggested higher mortality with PCI-DES among STEMI patients, with no statistically significant differences in SA or NSTEMI/UA. Intuitively, this makes sense, as STEMI patients most commonly undergo PCI of the culprit vessel and CABG is rarely utilized. Unfortunately, we did not assess complete revascularization post-STEMI because this preceded the COMPLETE trial.^35^

Our 5-year MACE findings are directionally consistent with prior evidence favoring CABG for MI prevention in diabetes and MVD. FREEDOM showed a lower 5-year risk of MACE with CABG (P=0.005), largely driven by fewer MI (P<0.001), but a higher stroke risk (P=0.03).^6^ In SYNTAX, the diabetes subgroup similarly showed higher 5-year MACCE (defined as all-cause mortality, cerebrovascular accident (CVA), MI or repeat revascularization) (P<0.001) after PCI-DES, largely driven by repeat revascularization, while MACE (P=0.26), all-cause death (P=0.065), stroke (P=0.34), or MI (P=0.20) did not differ significantly.^29^ We do believe CABG reduces repeat revascularization (and possibly MI), but this should be weighed against the lack of survival benefit at 5 years. This has implications for patients who may weigh these outcomes differently and should be fully informed before making a revascularization decision given the move towards a patient-centered decision-making model.

To our knowledge, a reversal of the mortality association beyond 5 years has not previously been described in a population-based cohort of patients with diabetes and MVD. In the restricted 10-year sub-cohort, PCI-DES was associated with lower all-cause mortality and cardiovascular mortality, but similar MACE compared to CABG. However, this modest association was not uniform across DJS strata, with the PCI-DES advantage most evident at lower DJS but higher mortality at a DJS of 10 and 12. Our 10-year estimates were based on a restricted sub-cohort in the early second-generation DES era, and long-term randomized data in this population have not shown a survival advantage for PCI-DES.^7^ The graded association with ischemic burden observed here has not previously been examined in a population-based diabetes cohort. While these findings are hypothesis-generating, they have implications for patients in whom 10-year survival after CABG is often a key consideration, yet for whom evidence remains sparse relative to second-generation DES PCI. We believe there are several reasons to support our findings. Graft failure at 5-10 years contributes to late risk after CABG and has been associated with recurrent ischemia and downstream adverse events during this time interval.^36^ Diabetes itself accelerates this process. Saphenous vein graft atherosclerosis appears to progress faster in patients with diabetes than in those without, which could compound attrition in conduit patency beyond 5 years in this population.^37,38^ New-onset postoperative atrial fibrillation, which complicates up to a third of CABG procedures, has independently been associated with higher long-term mortality, stroke, and renal failure, and may represent an additional pathway by which late risk accrues after CABG surgery.^39^ Unmeasured perioperative and postoperative factors could disadvantage CABG cumulatively over the following decade (e.g. cardiac surgery associated kidney injury, cerebral injury, Type V MI, etc.). While speculative, our data suggest that long-term mortality warrants further investigation.

Importantly, we have observed effect modification by anatomic complexity in other studies. Pooled patient-level evidence from SYNTAX, PRECOMBAT, and BEST reported effect modification by anatomic disease burden, suggesting a similar 5-year risk of all-cause mortality, MI, or stroke between PCI and CABG at lower-intermediate anatomic risk and worse outcomes with PCI at higher risk (P_interaction_=0.049).^40^ The 5-year interaction was no longer significant after excluding patients with left main disease, whereas the 10-year interaction remained, suggesting that jeopardized myocardium becomes relevant to strategy selection independently of left main disease only over extended follow-up.

Overall, key strengths include the large, diabetes-specific cohort and direct evaluation of effect modification by DJS when comparing PCI-DES and CABG. The real-world setting supports clinical relevance, and the near-complete 5-year follow-up minimizes loss-to-follow-up bias. Limitations include residual confounding and selection bias inherent to retrospective observational studies, and causal inference cannot be made. Patients who died between index catheterization and revascularization were not eligible for inclusion, and early deaths among patients destined for CABG may be underrepresented. Important determinants of treatment selection and prognosis, such as frailty and lifestyle factors, were unavailable. Comorbidities identified from administrative ICD-9/10 data may be under-ascertained.^41^ Detailed angiographic characteristics captured by the SYNTAX score were unavailable, although DJS is a validated proxy for anatomic disease burden.^42^ Residual confounding may persist despite IPTW because unmeasured determinants of treatment selection (e.g., frailty, surgical candidacy, and granular anatomy) may remain.^43^ Therefore, interpret the 5-year mortality findings as comparable long-term survival rather than equivalent mortality. The 10-year analysis was based on a smaller sub-cohort and should be interpreted as exploratory. Finally, as the study was conducted in a single-payer healthcare system, generalizability to other settings may be limited.

## Conclusion

Our study provides a real-world comparison of long-term outcomes between second-generation PCI-DES and CABG among patients with diabetes and MVD using IPTW-adjusted analyses. Overall, 5-year all-cause mortality was similar, whereas PCI-DES was associated with a higher risk of 5-year MACE. In an exploratory analysis of patients with potential 10-year follow-up, PCI-DES was associated with lower mortality, but similar MACE. The relative treatment effects varied by baseline anatomic complexity at both 5 and 10 years. Further studies are needed to clarify very long-term outcomes to better inform revascularization decision-making in patients with diabetes and MVD.

## Data Availability

The datasets used and analyzed during the current study are available from the corresponding author on reasonable request.

## Acknowledgements

This study is based in part on data provided by the PPHS (Primary & Preventative Health Services) and Alberta Health Services. The authors thank the Customer Relationship Management and Data Access Unit at PPHS for creating the linked database. The interpretation and conclusions are those of the researchers and do not represent the views of the Government of Alberta. Neither the Government of Alberta nor PPHS expressed any opinion in relation to this study. During the preparation of this work, the authors used ChatGPT version 5.2 to improve sentence formulation for clarity and readability. After using this tool, the authors reviewed and edited the content as needed and take full responsibility for the content of the publication. The datasets used and analyzed during the current study are available from the corresponding author on reasonable request.

## Nonstandard Abbreviations and Acronyms

CIF: Cumulative incidence function
DES: Drug-eluting stent
DJS: Duke Jeopardy Score
IPTW: Inverse probability of treatment weighting
MACCE: Major adverse cardiac and cerebrovascular events
MACE: Major adverse cardiovascular events
MVD: Multivessel coronary artery disease
PCI-DES: Percutaneous coronary intervention with drug-eluting stent
SYNTAX: Synergy between PCI with Taxus and Cardiac Surgery

## Supplemental Material

Tables S1–S5; Figures S1–S6

## References

1. Virani SS, Newby LK, Arnold SV, Bittner V, Brewer LC, Demeter SH, Dixon DL, Fearon WF, Hess B, Johnson HM, et al. 2023 AHA/ACC/ACCP/ASPC/NLA/PCNA Guideline for the Management of Patients With Chronic Coronary Disease: A Report of the American Heart Association/American College of Cardiology Joint Committee on Clinical Practice Guidelines. Circulation. 2023;148:e9–e119. doi: 10.1161/CIR.0000000000001168

2. Vrints C, Andreotti F, Koskinas KC, Rossello X, Adamo M, Ainslie J, Banning AP, Budaj A, Buechel RR, Chiariello GA, et al. 2024 ESC Guidelines for the management of chronic coronary syndromes. Eur Heart J. 2024;45:3415–3537. doi: 10.1093/eurheartj/ehae177

3. Byrne RA, Rossello X, Coughlan JJ, Barbato E, Berry C, Chieffo A, Claeys MJ, Dan GA, Dweck MR, Galbraith M, et al. 2023 ESC Guidelines for the management of acute coronary syndromes. Eur Heart J. 2023;44:3720–3826. doi: 10.1093/eurheartj/ehad191

4. Rao SV, O’Donoghue ML, Ruel M, Rab T, Tamis-Holland JE, Alexander JH, Baber U, Baker H, Cohen MG, Cruz-Ruiz M, et al. 2025 ACC/AHA/ACEP/NAEMSP/SCAI Guideline for the Management of Patients With Acute Coronary Syndromes: A Report of the American College of Cardiology/American Heart Association Joint Committee on Clinical Practice Guidelines. Circulation. 2025;151:e771–e862. doi: 10.1161/CIR.0000000000001309

5. Lawton JS, Tamis-Holland JE, Bangalore S, Bates ER, Beckie TM, Bischoff JM, Bittl JA, Cohen MG, DiMaio JM, Don CW, et al. 2021 ACC/AHA/SCAI Guideline for Coronary Artery Revascularization: A Report of the American College of Cardiology/American Heart Association Joint Committee on Clinical Practice Guidelines. Circulation. 2022;145:e18–e114. doi: 10.1161/CIR.0000000000001038

6. Farkouh ME, Domanski M, Sleeper LA, Siami FS, Dangas G, Mack M, Yang M, Cohen DJ, Rosenberg Y, Solomon SD, et al. Strategies for multivessel revascularization in patients with diabetes. N Engl J Med. 2012;367:2375–2384. doi: 10.1056/NEJMoa1211585

7. Farkouh ME, Domanski M, Dangas GD, Godoy LC, Mack MJ, Siami FS, Hamza TH, Shah B, Stefanini GG, Sidhu MS, et al; FREEDOM Follow-On Study Investigators. Long-term survival following multivessel revascularization in patients with diabetes: the FREEDOM Follow-On Study. J Am Coll Cardiol. 2019;73:629–638. doi: 10.1016/j.jacc.2018.11.001

8. Hannan EL, Wu C, Walford G, Culliford AT, Gold JP, Smith CR, Higgins RSD, Carlson RE, Jones RH. Drug-eluting stents vs. coronary-artery bypass grafting in multivessel coronary disease. N Engl J Med. 2008;358:331–341. doi: 10.1056/NEJMoa071804

9. Kim YG, Park DW, Lee WS, Park GM, Sun BJ, Lee CH, Hwang KW, Cho SW, Kim YR, Song HG, et al. Influence of diabetes mellitus on long-term (five-year) outcomes of drug-eluting stents and coronary artery bypass grafting for multivessel coronary revascularization. Am J Cardiol. 2012;109:1548–1557. doi: 10.1016/j.amjcard.2012.01.377

10. Bangalore S, Toklu B, Feit F. Outcomes with coronary artery bypass graft surgery versus percutaneous coronary intervention for patients with diabetes mellitus: can newer generation drug-eluting stents bridge the gap? Circ Cardiovasc Interv. 2014;7:518–525. doi: 10.1161/CIRCINTERVENTIONS.114.001346

11. Bangalore S, Guo Y, Samadashvili Z, Blecker S, Xu J, Hannan EL. Everolimus eluting stents vs. coronary artery bypass graft surgery for patients with diabetes and multivessel disease. Circ Cardiovasc Interv. 2015;8:e002626. doi: 10.1161/CIRCINTERVENTIONS.115.002626

12. Califf RM, Phillips HR, Hindman MC, Mark DB, Lee KL, Behar VS, Johnson RA, Pryor DB, Rosati RA, Wagner GS. Prognostic value of a coronary artery jeopardy score. J Am Coll Cardiol. 1985;5:1055–1063. doi: 10.1016/s0735-1097(85)80005-x

13. Public Health Agency of Canada. The Canadian Chronic Disease Surveillance System – An Overview. Available at https://www.canada.ca/en/public-health/services/publications/canadian-chronic-disease-surveillance-system-factsheet.html. Accessed November 27, 2025.

14. Ghali WA, Knudtson ML. Overview of the Alberta Provincial Project for Outcome Assessment in Coronary Heart Disease. On behalf of the APPROACH investigators. Can J Cardiol. 2000;16:1225–1230.

15. Quan H, Sundararajan V, Halfon P, Fong A, Burnand B, Luthi JC, Saunders LD, Beck CA, Feasby TE, Ghali WA. Coding algorithms for defining comorbidities in ICD-9-CM and ICD-10 administrative data. Med Care. 2005;43:1130–1139. doi: 10.1097/01.mlr.0000182534.19832.83

16. Tonelli M, Wiebe N, Fortin M, Guthrie B, Hemmelgarn BR, James MT, Klarenbach SW, Lewanczuk R, Manns BJ, Ronksley P, et al. Methods for identifying 30 chronic conditions: application to administrative data. BMC Med Inform Decis Mak. 2015;15:31. doi: 10.1186/s12911-015-0155-5

17. Government of Alberta. Official Standard Geographic Areas. 2018. Available at https://open.alberta.ca/publications/official-standard-geographic-areas. Accessed April 27, 2025.

18. Pampalon R, Hamel D, Gamache P, Raymond G. A deprivation index for health planning in Canada. Chronic Dis Can. 2009;29:178–191.

19. Statistics Canada. Dissemination area (DA) – Census Dictionary. Available at https://www12.statcan.gc.ca/census-recensement/2011/ref/dict/geo021-eng.cfm. Accessed August 26, 2025.

20. Cummins C, Winter H, Cheng KK, Maric R, Silcocks P, Varghese C. An assessment of the Nam Pehchan computer program for the identification of names of south Asian ethnic origin. J Public Health Med. 1999;21:401–406. doi: 10.1093/pubmed/21.4.401

21. Quan H, Wang F, Schopflocher D, Norris C, Galbraith PD, Faris P, Graham MM, Knudtson ML, Ghali WA. Development and validation of a surname list to define Chinese ethnicity. Med Care. 2006;44:328–333. doi: 10.1097/01.mlr.0000204010.81331.a9

22. Shah BR, Chiu M, Amin S, Ramani M, Sadry S, Tu JV. Surname lists to identify South Asian and Chinese ethnicity from secondary data in Ontario, Canada: a validation study. BMC Med Res Methodol. 2010;10:42. doi: 10.1186/1471-2288-10-42

23. Statistics Canada. Census Profile, 2021 Census of Population. Available at https://www12.statcan.gc.ca/census-recensement/2021/dp-pd/prof/index.cfm?Lang=E. Accessed August 24, 2026.

24. Taggart DP. Percutaneous coronary interventions versus coronary artery bypass graft surgery in coronary artery disease. Vascul Pharmacol. 2024;155:107367. doi: 10.1016/j.vph.2024.107367

25. Lee YJ, Hong SJ, Kim BK, Shin S, Suh Y, Kim S, Ahn CM, Kim JS, Ko YG, Choi D, et al. Long-term outcomes after percutaneous coronary intervention relative to bypass surgery in diabetic patients with multivessel coronary artery disease according to clinical presentation. Coron Artery Dis. 2020;31:174–183. doi: 10.1097/MCA.0000000000000767

26. Haukoos JS, Newgard CD. Advanced statistics: missing data in clinical research—part 1: an introduction and conceptual framework. Acad Emerg Med. 2007;14:662–668. doi: 10.1111/j.1553-2712.2007.tb01855.x

27. Austin PC. Balance diagnostics for comparing the distribution of baseline covariates between treatment groups in propensity-score matched samples. Stat Med. 2009;28:3083–3107. doi: 10.1002/sim.3697

28. Austin PC. The performance of different propensity score methods for estimating marginal hazard ratios. Stat Med. 2013;32:2837–2849. doi: 10.1002/sim.5705

29. Kappetein AP, Head SJ, Morice MC, Banning AP, Serruys PW, Mohr FW, Dawkins KD, Mack MJ; SYNTAX Investigators. Treatment of complex coronary artery disease in patients with diabetes: 5-year results comparing outcomes of bypass surgery and percutaneous coronary intervention in the SYNTAX trial. Eur J Cardiothorac Surg. 2013;43:1006–1013. doi: 10.1093/ejcts/ezt017

30. Di Gioia G, Soto Flores N, Franco D, Colaiori I, Sonck J, Gigante C, Kodeboina M, Bartunek J, Vanderheyden M, Van Praet F, et al. Coronary artery bypass grafting or fractional flow reserve-guided percutaneous coronary intervention in diabetic patients with multivessel disease. Circ Cardiovasc Interv. 2020;13:e009157. doi: 10.1161/CIRCINTERVENTIONS.120.009157

31. Navarese EP, Kowalewski M, Kandzari D, Lansky A, Górny B, Kołtowski Ł, Waksman R, Berti S, Musumeci G, Limbruno U, et al. First-generation versus second-generation drug-eluting stents in current clinical practice: updated evidence from a comprehensive meta-analysis of randomised clinical trials comprising 31 379 patients. Open Heart. 2014;1:e000064. doi: 10.1136/openhrt-2014-000064

32. Bangalore S, Toklu B, Patel N, Feit F, Stone GW. Newer-generation ultrathin strut drug-eluting stents versus older second-generation thicker strut drug-eluting stents for coronary artery disease. Circulation. 2018;138:2216–2226. doi: 10.1161/CIRCULATIONAHA.118.034456

33. Bansilal S, Bonaca MP, Cornel JH, Storey RF, Bhatt DL, Steg PG, Im K, Murphy SA, Angiolillo DJ, Kiss RG, et al. Ticagrelor for secondary prevention of atherothrombotic events in patients with multivessel coronary disease. J Am Coll Cardiol. 2018;71:489–496. doi: 10.1016/j.jacc.2017.11.050

34. Zinman B, Wanner C, Lachin JM, Fitchett D, Bluhmki E, Hantel S, Mattheus M, Devins T, Johansen OE, Woerle HJ, et al. Empagliflozin, cardiovascular outcomes, and mortality in type 2 diabetes. N Engl J Med. 2015;373:2117–2128. doi: 10.1056/NEJMoa1504720

35. Mehta SR, Wood DA, Storey RF, Mehran R, Bainey KR, Nguyen H, Meeks B, Di Pasquale G, López-Sendón J, Faxon DP, et al. Complete revascularization with multivessel PCI for myocardial infarction. N Engl J Med. 2019;381:1411–1421. doi: 10.1056/NEJMoa1907775

36. Gaudino M, Antoniades C, Benedetto U, Deb S, Di Franco A, Di Giammarco G, Fremes S, Glineur D, Grau J, He GW, et al. Mechanisms, consequences, and prevention of coronary graft failure. Circulation. 2017;136:1749–1764. doi: 10.1161/CIRCULATIONAHA.117.027597

37. Sun Y, Kang L, Li J, Liu H, Wang Y, Wang C, Zou Y. Advanced glycation end products impair the functions of saphenous vein but not thoracic artery smooth muscle cells through RAGE/MAPK signalling pathway in diabetes. J Cell Mol Med. 2016;20:1945–1955. doi: 10.1111/jcmm.12886

38. Akhrass R, Bakaeen FG. The advantage of surgical revascularization in diabetic patients with multivessel disease: more arterial conduits, more benefit. J Thorac Cardiovasc Surg. 2022;164:119–122. doi: 10.1016/j.jtcvs.2021.01.140

39. El-Chami MF, Kilgo P, Thourani V, Lattouf OM, Delurgio DB, Guyton RA, Leon AR, Puskas JD. New-onset atrial fibrillation predicts long-term mortality after coronary artery bypass graft. J Am Coll Cardiol. 2010;55:1370–1376. doi: 10.1016/j.jacc.2009.10.058

40. Cavalcante R, Sotomi Y, Mancone M, Lee CW, Ahn JM, Onuma Y, Lemos PA, van Geuns RJ, Park SJ, Serruys PW. Impact of the SYNTAX scores I and II in patients with diabetes and multivessel coronary disease: a pooled analysis of patient level data from the SYNTAX, PRECOMBAT, and BEST trials. Eur Heart J. 2017;38:1969–1977. doi: 10.1093/eurheartj/ehx138

41. Quan H, Li B, Saunders LD, Parsons GA, Nilsson CI, Alibhai A, Ghali WA; IMECCHI Investigators. Assessing validity of ICD-9-CM and ICD-10 administrative data in recording clinical conditions in a unique dually coded database. Health Serv Res. 2008;43:1424–1441. doi: 10.1111/j.1475-6773.2007.00822.x

42. Graham MM, Faris PD, Ghali WA, Galbraith PD, Norris CM, Badry JT, Mitchell LB, Curtis MJ, Knudtson ML. Validation of three myocardial jeopardy scores in a population-based cardiac catheterization cohort. Am Heart J. 2001;142:254–262. doi: 10.1067/mhj.2001.116481

43. Austin PC. An introduction to propensity score methods for reducing the effects of confounding in observational studies. Multivariate Behav Res. 2011;46:399–424. doi: 10.1080/00273171.2011.568786

